# Intensity of care and the health status of caregivers to elderly rural South Africans

**DOI:** 10.1101/2024.10.16.24315588

**Authors:** Sostina S Matina, Lenore Manderson, F. Xavier Gómez-Olivé, Lisa Berkman, Guy Harling

## Abstract

**Objectives:** Informal caregivers play an indispensable role in and are often the sole source of care for older adults in low and middle-income settings worldwide. Intensive informal care predicts mortality and morbidity among caregivers in higher-income settings. However, there is limited evidence from poorer settings, including Africa countries, where caregiving is shared widely, including across generations. We therefore investigated caregivers’ health status in rural South Africa.

**Methods:** We conducted quantitative interviews with all household members and all non-household caregivers aged ≥12 (n=1012) of 106 older adults in rural Mpumalanga, South Africa. We used multivariable regression with care-recipient random intercepts to assess the relationships between four caregiving characteristics and both self-reported chronic conditions and self-reported health status, considering how caregiver age moderated each association.

**Results:** Over half of all caregivers reported at least one chronic health condition, despite half being aged under 40. Caregivers self-reporting the worst health status provided high hours of care. However, caregivers’ health status was not significantly associated with weekly care quantity or history of caring. Those aged ≥40 who reported being a main caregiver had 52% increased odds of reporting poorer health compared to other same-aged carers (95% confidence interval: 0.99, 2.35), while having more chronic conditions was associated with being expected to act as a sole caregiver more often among caregivers aged ≤39.

**Discussion:** Greater caring responsibilities for older adults were not consistently associated with caregivers’ health in a setting where poor health is common, and caregiving is spread widely. Longitudinal data is necessary to unpack possible explanations of these findings, and to determine whether intensive caregiving speeds downward health trajectories for carers.

## INTRODUCTION

As the populations of lower- and middle-income countries (LMICs) age, multimorbidity is becoming a key challenge associated with adverse health outcomes, including reduced physical function and quality of life, and for governments, the cost of meeting chronic healthcare needs (1, 2). Multimorbidity also increases health facility demand and the risk of sub-optimal care due to conflicting advice on treatment and care and the difficulty of managing multiple medications (3–5). At the same time that there is a high mortality from infectious diseases in South Africa, long-term conditions are associated with increasing morbidity and mortality, accounting for 66% of the disease burden in men and 77% in women (2, 6). The prevalence of multimorbidity is more pronounced in rural areas than urban areas (7, 8). As this population ages and the number of conditions increases, there is likely a need for increased care. This is usually provided informally (9, 10): there are limited options for institutional formal care in LMICs (11–14), even if this were considered desirable.

Informal caregiving is ubiquitous in Sub-Saharan Africa (SSA) (9, 15, 16) and all LMICs, and is mostly provided by family members of care recipients (17). Family caregivers play an indispensable role and are often the sole caregivers for older family members (13, 18). These caregiving roles extend beyond assistance with daily activities to encompass emotional, social and financial support, medication management, and accompanying care recipients to health facilities (12, 16). In rural settings where access to formal healthcare services may be limited, family caregivers are primarily responsible for managing the diverse needs of elderly or otherwise frail family members (16, 19). Most rural areas in SSA are characterised by multigenerational families and close-knit communities, whose members may contribute to shared caregiving responsibilities (12, 16, 17, 20). Comparatively, caregivers in high-income countries (HICs) often share responsibilities with other formal caregivers and receive substantial support from healthcare professionals and community services (21). The lack of such formal support in LMIC, including SSA, may result in primary caregivers bearing the full burden of care, resulting in significant physical and emotional stress.

Providing intensive informal care to older adults is reported as an independent risk factor for higher mortality (22–24) and morbidity (25, 26). Caregivers of people living with cognitive impairment report a more significant subjective burden and lower quality of life than other caregivers (27). Caregivers in HICs who provide more than 20 hours of care a week and those who have spent more years giving care are more likely to report mental distress, suggesting that intensive caregiving is a risk factor for ill health. Studies in HIC further report that caregivers over 40 years of age are more likely than younger caregivers to report a cognitive decline and adverse physical health (14, 25, 28). However, there is a notable dearth of literature on the impact of the intensity of caregiving on caregivers’ health in LMICs.

Care recipient expectations encompass a broad spectrum of needs, including physical assistance, emotional support, empathy, and respect for autonomy (29). Meeting these expectations not only enhances the well-being and satisfaction of the care recipient but also has significant implications for their health outcomes (14). Conversely, unmet expectations can lead to dissatisfaction, stress, and a decline in both mental and physical health (30). On the other hand, prolonged caregiving is associated with chronic stress and strain, which can also lead to a range of adverse health outcomes. These include physical ailments such as musculoskeletal problems and cardiovascular diseases, as well as mental health issues like depression, anxiety, and caregiver burnout (22).

While some studies indicate that an increase in caregiving hours is directly linked to greater burden, the care recipient’s condition – particularly cognitive impairments like dementia – plays a more significant role in shaping emotional and physical strain. As cognitive abilities decline, caregivers face an ongoing sense of grief and loss, often referred to as ambiguous loss, where the person is physically present but psychologically absent (31). This form of loss is particularly distressing because it is prolonged and unresolved, leaving caregivers in a state of continuous mourning. Caregivers adapt to the loss of shared memories, emotional connections, and basic communication, leading to isolation and significant emotional strain (32). This emotional toll is compounded by role reversal, where caregivers feel they are losing a spouse, parent, or partner, despite their physical presence, amplifying feelings of grief and emotional exhaustion (33). As a result, caregivers of individuals with dementia or Alzheimer’s often experience higher rates of depression, anxiety, and psychological distress compared to those caring for people with other conditions (34, 35).

In HICs, interventions supporting caregivers have improved caregivers’ health outcomes, especially their mental health (24, 26, 36). For instance, Resources for Enhancing Alzheimer’s Caregiver Health Intervention (REACH) in the USA improved caregiver health outcomes, including self-reported health, physical health, and emotional health (36). Through a nuanced understanding of the caregiver-care recipient relationship and its impact on health in a setting with an already high prevalence of chronic conditions (37, 38), public health professionals and healthcare practitioners can develop targeted interventions that prioritise the wellbeing of both caregivers and care recipients.

This study investigated caregivers’ health status (chronic conditions and self-reported health) in rural South Africa. This rural population has limited access to formal care for support or respite care, and plays a significant role in the informal caregiving of older, frail family members (39).

## METHODS

### Setting

Our study was nested within the Agincourt Health and Socio-Demographic Surveillance System (HDSS) site of the South African Medical Research Council/University of the Witwatersrand Rural Public Health and Health Transitions Research Unit, in Bushbuckridge, Mpumalanga Province, South Africa (37). This region is marked by constrained employment opportunities and limited access to formal care services, intensified by the presence of multimorbidity (37, 38). This setting provides a context for examining the intricate interplay between caregiving responsibilities, socioeconomic challenges, and health outcomes.

### Study design

#### Participant selection and sampling strategy

Our study, Kaya (“home” in the local xi-Tsonga language), aimed to understand how care is provided to older adults with substantial care needs in the Agincourt HDSS site (40). To achieve this, we sampled 116 index cases (older people with or at risk of cognitive impairment, hereafter “care recipients”). These care recipients were sampled from the Health and Aging in Africa: A Longitudinal Study of an INDEPTH Community in South Africa (HAALSI) study, a longitudinal population-representative sample of HDSS residents aged ≥40 established in 2014 (38). The HAALSI Dementia study began in 2019-20 to estimate the prevalence and incidence of dementia and mild cognitive impairment in the HAALSI cohort, based on a risk-stratified HAALSI subsample, with a second round in 2021 (41). We sampled individuals with the highest predicted dementia severity in the Dementia Study, stratified to include an equal number of male and female care recipients. Further details on our sampling process are available elsewhere (39).

We first interviewed an index household respondent (“primary respondent”) for each care recipient. We asked the index person to list all resident and non-resident household members, plus any non-household kin or non-kin who provided care to the care recipient. Quantitative interviews were conducted with all household members, and all non-household caregivers aged ≥12. Data were collected using face-to-face interviews between July and December 2022 at participants’ homes. Interviewers used tablet computers to capture data.

### Ethical considerations

The study received ethics approval from the University of the Witwatersrand Human Research Ethics Committee (REC) (Medical; M200373), University College London REC (152311/001) and Mpumalanga Province Health Research Committee, and letters of support from the Mpumalanga Department of Health and the Agincourt Community Advisory Board. All adult participants provided written informed consent; all minors provided written informed assent following parent/guardian written consent.

### Measures

We assessed caregiver health using two key measures of poor health. First, we used self-reported current health status on a five-point scale (very good, good, moderate, bad, very bad) with worst health as the highest category. Second, we used self-reports of any past diagnosis of a range of chronic conditions and depressive symptoms: type II diabetes; asthma; kidney disease; cancer; cardiovascular disease (hypertension, stroke, angina, heart attack or hypercholesterolemia); HIV; tuberculosis; degenerative brain disorders (Parkinson’s, Alzheimer’s, non-specific memory problems); musculoskeletal conditions; and any other condition (headaches, chronic disease, serious illness and ulcers). Caregivers also completed the Center for Epidemiologic Studies Depression 10-item scale (CESD), for which we considered a score of ≥10 as indicative of being at risk for clinical depression (42). We generated a count of conditions reported, and defined multimorbidity as a count of two or more (43, 44).

We calculated four measures of caring level. We measured both average weekly hours of care provided (continuous and binary at ≥20 hours) and length of care provided in years. We also considered whether caregivers self-reported that they were the “main caregiver” and the frequency with which they agreed that the care recipient “expects you to care as only dependable person” (never, rarely, sometimes, quite frequently and nearly always).

We considered as potential confounders: age (<18, 19-39, 40-59, ≥60); gender; marital status (married/coresident, never married, previously married); household membership; education (none, any primary, any secondary, any tertiary); and work status (fulltime work, part-time work, seeking work, out of workforce – either not seeking work at present or retired).

### Statistical analyses

We generated descriptive statistics using median and interquartile range (IQR) for continuous variables and percentages for categorical variables. We assessed differences in exposures and outcomes by age group using Pearson’s X^2^ tests for categorical variables and Wilcoxon rank-sum tests for continuous variables. We used UpSet plots to visualise the distribution and co-occurrence of self-reported chronic conditions (45, 46). We used multivariable regression, including random intercepts for care recipient identity, to assess the relationships between self-reported chronic condition count (Poisson models) and self-reported health status (ordinal logistic models) and the four caregiving characteristics. We included all potential confounders as covariates. We first ran eight separate models using one outcome and one care variable each, and then one model per outcome, including all care variables. We used R version 4.3.2 for our statistical analyses (R Development Core Team, 2010).

## RESULTS

Of the 116 sampled care recipients, two had passed away, four were unlocatable, and in four households, the primary respondents declined to participate. A primary respondent consented and was interviewed among the remaining 106 care recipients (55 female, 51 male) living in 24 villages. Among these 106 recipients, 19 were predicted to have moderate/severe dementia, 71 to have mild dementia and 16 no dementia. Almost all named household and non-household members who provided care to the recipients (1012/1020, 99.2%) consented and completed a survey (three outmigrated, one died, three non-contactable, one declined). Three-fifths of caregivers were female, with most either aged 19-39 (38.0%) or 40-59 (30.6%) (Table 1). Only a minority (29.4%) were employed, and a majority were either non-household relatives (51.2%) or non-relatives (14.3%).

**Table 1:** Caregiver socio-demographic and health status characteristics.

| Characteristic | Overall,<br>N = 1,012<sup>1</sup> | Under 18,<br>N = 140 | 19-39,<br>N = 385 | 40-59,<br>N = 310 | 60+,<br>N = 177 | p-value<sup>2</sup> |
| --- | --- | --- | --- | --- | --- | --- |
| Female sex, % | 60.8% | 60.0% | 54.8% | 64.5% | 67.8% | 0.01 |
| Marital status, % |  |  |  |  |  | <0.001 |
| Married/coresident | 36.5% | 1.4% | 28.6% | 56.8% | 45.8% |  |
| Never married | 42.5% | 98.6% | 65.7% | 11.6% | 1.7% |  |
| Previously married | 21.0% | 0.0% | 5.7% | 31.6% | 52.5% |  |
| Work status, % |  |  |  |  |  | <0.001 |
| Fulltime work | 19.8% | 0.0% | 25.2% | 31.3% | 3.4% |  |
| Part-time work | 9.6% | 0.0% | 11.4% | 16.5% | 1.1% |  |
| Seeking work | 22.5% | 3.6% | 37.1% | 24.2% | 2.8% |  |
| Out of workforce | 48.1% | 96.4% | 26.2% | 28.1% | 92.7% |  |
| Relationship type, % |  |  |  |  |  |  |
| Spouse | 3.2% | 0.0% | 0.0% | 2.3% | 14.2% |  |
| Household Relative | 31.3% | 51.4% | 44.4% | 20.6% | 5.1% |  |
| Non-household Relative | 51.2% | 45.0% | 44.4% | 61.6% | 52.8% |  |
| Non-relative | 14.3% | 3.6% | 11.2% | 15.5% | 27.8% |  |
| Education, % |  |  |  |  |  | <0.001 |
| None | 13.5% | 0.0% | 0.5% | 11.6% | 55.9% |  |
| Primary Education | 19.8% | 26.4% | 6.5% | 23.2% | 37.3% |  |
| Secondary Education | 56.5% | 72.9% | 78.2% | 51.9% | 4.5% |  |
| Tertiary Education | 10.2% | 0.7% | 14.8% | 13.2% | 2.3% |  |
| Caregiving (self-reported) |  |  |  |  |  |  |
| Main caregiver | 29.2% | 17.9% | 27.3% | 32.6% | 36.3% |  |
| Length of care (years) | 2 (1, 7) | 4 (1, 7) | 2 (1, 6) | 2 (1, 6) | 3 (1, 8) | 0.02 |
| Weekly care provision (hours) | 3.0 (0.6, 7) | 2.0 (0.6, 4) | 3.0 (0.5, 5) | 4.0 (0.4, 10) | 5.0 (2, 10) | <0.001 |
| ≥20 hours care per week | 5.4% | 2.9% | 5.5% | 4.5% | 9.1% | 0.07 |
| Expected to be only dependable caregiver |  |  |  |  |  | 0.05 |
| Never | 54.7% | 50.7% | 55.6% | 52.9% | 59.3% |  |
| Rarely | 22.0% | 25.7% | 23.4% | 21.6% | 16.9% |  |
| Sometimes | 15.4% | 15.7% | 16.9% | 14.2% | 14.1% |  |
| Quite frequently | 6.1% | 7.1% | 3.4% | 8.7% | 6.8% |  |
| Nearly always | 1.7% | 0.7% | 0.8% | 2.6% | 2.8% |  |
| Health Conditions |  |  |  |  |  |  |
| Kidney disease, % | 0.2% | 0.0% | 0.3% | 0.0% | 0.6% | 0.60 |
| Cognitive decline, % | 0.4% | 0.0% | 0.0% | 0.3% | 1.7% | 0.02 |
| Depression symptoms, % | 19.2% | 12.9% | 19.0% | 21.3% | 20.9% | 0.2 |
| HIV seropositive, % | 7.9% | 0.7% | 6.0% | 14.8% | 5.6% | <0.001 |
| Diabetes, % | 3.2% | 0.0% | 0.5% | 2.6% | 12.4% | <0.001 |
| Musculoskeletal, % | 1.2% | 0.0% | 0.8% | 1.3% | 2.8% | 0.11 |
| Tuberculosis, % | 0.8% | 0.0% | 0.8% | 0.3% | 2.3% | 0.11 |
| Cancer, % | 0.3% | 0.0% | 0.0% | 0.6% | 0.6% | 0.30 |
| Other conditions, % | 0.5% | 0.0% | 0.5% | 0.6% | 0.6% | 0.98 |
| CVDs, % | 25.7% | 2.9% | 9.9% | 31.6% | 67.8% | <0.001 |
| Count of conditions, % |  |  |  |  |  |  |
| 0 | 56.6% | 84.3% | 71.7% | 46.1% | 20.3% |  |
| 1 | 30.0% | 15.0% | 20.0% | 37.7% | 50.3% |  |
| 2 | 10.9% | 0.7% | 7.3% | 12.9% | 23.2% |  |
| 3 | 2.4% | 0.0% | 1.0% | 2.9% | 6.2% |  |
| 4 | 0.1% | 0.0% | 0.0% | 0.3% | 0.0% |  |
| Multimorbidity, % | 13.3% | 0.7% | 8.3% | 16.1% | 29.4% | <0.001 |
| Self-reported health |  |  |  |  |  |  |
| Very good | 41.0% | 69.3% | 50.9% | 33.2% | 10.7% |  |
| Good | 43.5% | 25.7% | 40.0% | 53.5% | 47.5% |  |
| Moderate | 13.1% | 5.0% | 7.5% | 11.3% | 35.0% |  |
| Bad | 2.0% | 0.0% | 0.5% | 1.9% | 6.8% |  |
| Very bad | 0.4% | 0.0% | 1.0% | 0.0% | 0.0% |  |

<sup>1</sup>%; Median (IQR), <sup>2</sup>Pearson's Chi-squared test; Kruskal-Wallis rank sum test; Fisher's exact test.1 missing for weekly hours

Almost all individuals nominated as caregivers (96.6%) reported giving care; 29.2% of them reported that they were the main caregiver, being higher for caregivers aged over 60 (36.3%). The median reported length of care was two years (IQR: 1.7), with the greatest length among non-working people, including those aged under 18 and those over 60. Caregivers provided a median of three hours of care weekly (IQR: 0.6.7), a quantity that increased with caregiver’s age. Only 5.4% of caregivers provided 20 hours or more of weekly care; this higher level of care was again most common in the caregivers over 60. Around half of caregivers (54.7%) reported that they were never expected to be the only dependable caregiver, while around one in 13 individuals (7.8%) were expected to play this role quite frequently or nearly always, with slight variation by the age of the caregiver.

Almost half (45.4%) of caregivers reported a chronic health condition, with hypertension, depressive symptoms and HIV being the most common (Table 1). Multimorbidity was substantially less common, with 13.3% of caregivers reporting two or more conditions, although this figure rose to 29.4% above age 60. The most common condition combinations were hypertension and depressive symptoms, followed by hypertension and diabetes (Figure 1). Participants with three chronic conditions were most likely to report hypertension, depressive symptoms, and HIV. Most caregivers reported their health as very good (41%) or good (43%), with higher values at younger ages. Although almost no caregiver under 39 years of age reported bad or very bad health (1%), 13% of caregivers under 18 reported symptoms suggestive of depression.

**Figure 1:**
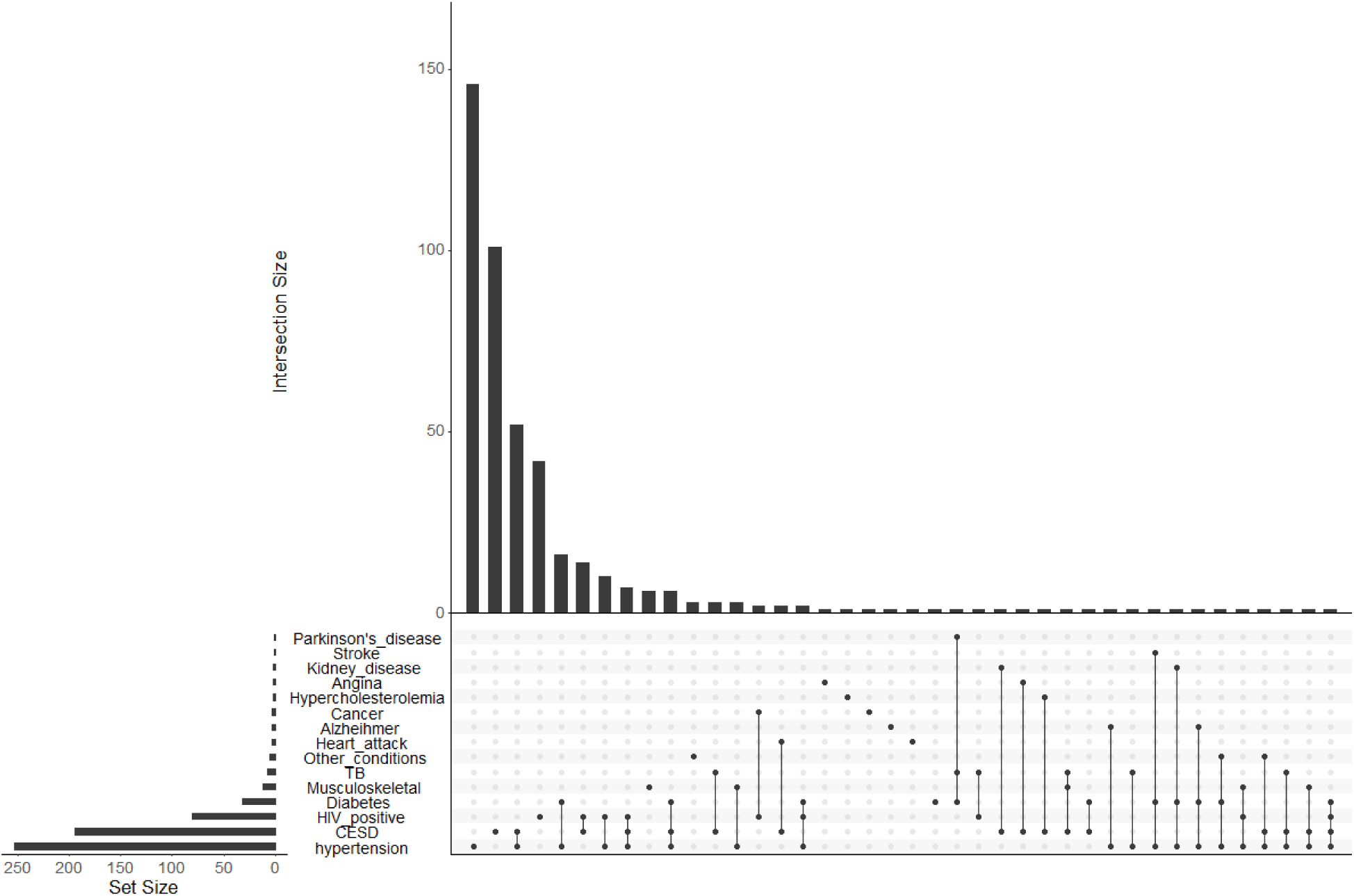
Prevalence and combinations of chronic conditions reported by caregivers.

In bivariate analyses, the number of reported chronic conditions by the caregivers was not significantly associated with weekly hours of care provided (Figure 2), although caregivers with three or more conditions provided somewhat less care (mean 4.0 hours) versus 6.0 hours (for two conditions), 5.6 hours (for one condition) and 5.4 hours (for no chronic condition). In contrast, caregivers who reported “very bad” health (mean 11.2 hours) or “bad” health (mean 9.2 hours) reported more hours providing weekly care compared to those with “very good” (mean 5.3 hours), “good” (mean 5.3 hours), “or “moderate” health (mean 5.9 hours).

**Figure 2:**
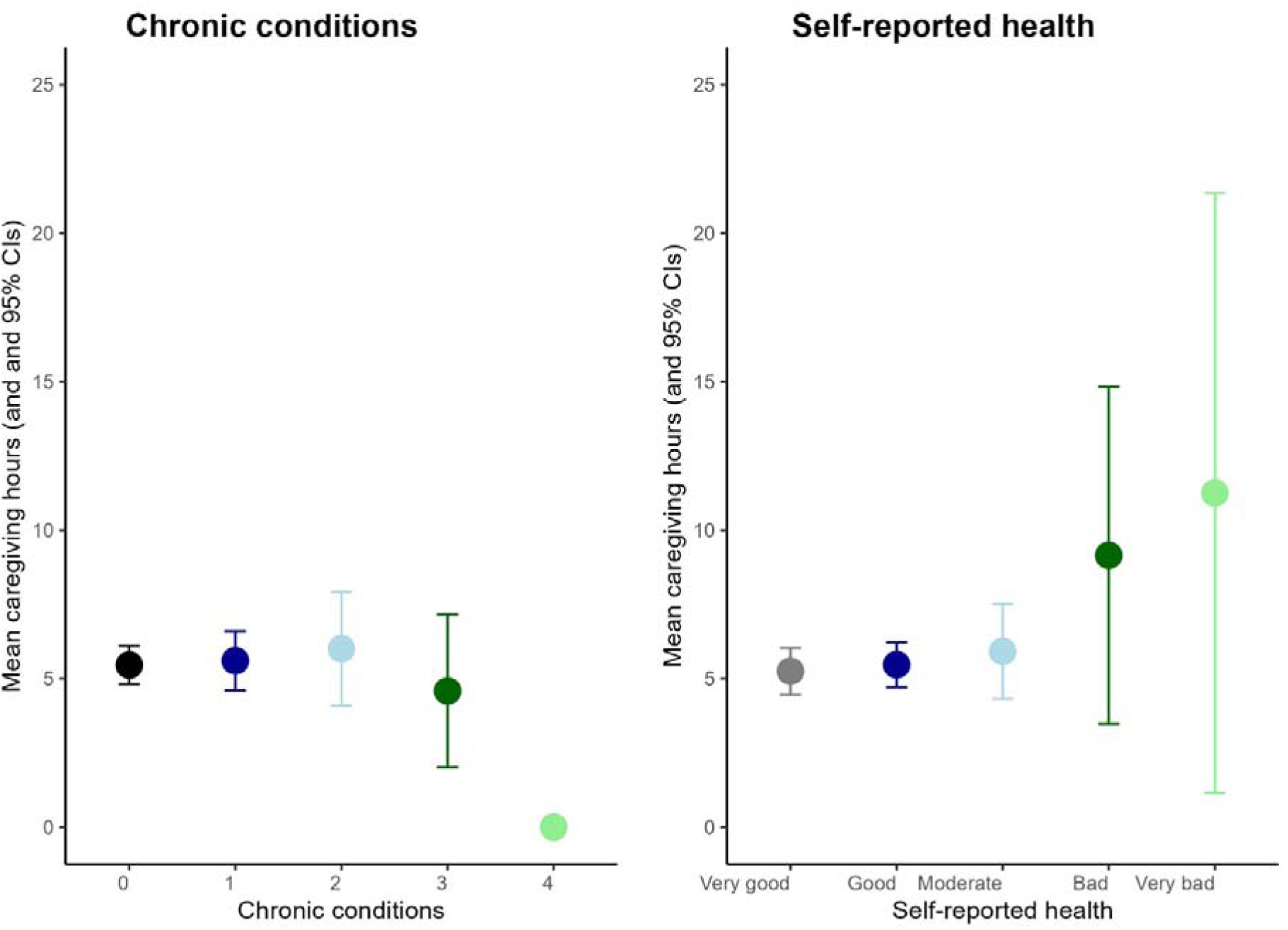
Bivariate association between weekly hours of care and: (A) chronic health conditions; and (B) self-reported health.

In multivariable models, the association between the number of health conditions and expectations of care was inverse U-shaped, with the greatest health burden amongst those caregivers ‘sometimes’ expected by the care recipient to be the only dependable caregiver (prevalence ratio [PR] 1.50, 95%CI 1.18-1.94 vs those never expected to care), and least ill-health amongst those ‘rarely’, ‘quite frequently’ or ‘nearly always’ expected to play this role (Table 2). This pattern was unchanged in a stratified model for caregivers aged ≥40. However, among younger caregivers, the association was uniformly positive, with those expected to care ‘quite frequently/almost always’ most likely to report chronic conditions (PR: 2.00, 95%CI: 1.03-3.91 vs. never). Other predictors of reporting more chronic conditions were limited among all participants and for older caregivers, but caregivers aged ≤39 with more chronic conditions were slightly more likely to report being a main caregiver and had been providing care for longer but less likely to provide intensive care now.

**Table 2:** Multivariable regression models of self-reported health and care provision.

|  | Number of chronic conditions |  | Worse self-reported quality of health |  |
| --- | --- | --- | --- | --- |
|  | Single care variable | All care variables | Single care variable | All care variables |
| **PANEL 1: ALL PARTICIPANTS** |  |  |  |  |
| CR expects you to care as the only dependable person (vs Never) |  |  |  |  |
| Rarely | 1.34 [1.07, 1.68] | 1.33 [1.05, 1.68] | 1.28 [0.94, 1.75] | 1.36 [0.99, 1.88] |
| Sometimes | 1.49 [1.17, 1.88] | 1.50 [1.18, 1.92] | 1.47 [1.04, 2.10] | 1.40 [0.97, 2.01] |
| Quite frequently | 1.33 [0.96, 1.84] | 1.34 [0.96, 1.89] | 1.21 [0.71, 2.06] | 1.10 [0.64, 1.91] |
| Nearly always | 1.20 [0.65, 2.20] | 1.19 [0.64, 2.25] | 1.03 [0.38, 2.83] | 0.79 [0.28, 2.20] |
| ≥ 20 weekly hours of care (binary) | 1.18 [0.83, 1.67] | 1.02 [0.69, 1.49] | 1.41 [0.82, 2.42] | 1.11 [0.62, 1.97] |
| Length of care (years) | 1.07 [0.93, 1.22] | 1.04 [0.90, 1.20] | 1.16 [0.95, 1.42] | 1.08 [0.87, 1.35] |
| Self-reported main caregiver | 1.11 [0.91, 1.36] | 1.04 [0.84, 1.29] | 1.46 [1.11, 1.92] | 1.44 [1.06, 1.94] |
| **PANEL 2: AGE UNDER 40** |  |  |  |  |
| CR expects you to care as the only dependable person (vs Never) |  |  |  |  |
| Rarely | 1.34 [1.05, 2.55] | 1.81 [1.15, 2.84] | 1.07 [0.90, 1.27] | 1.08 [0.90, 1.27] |
| Sometimes | 1.48 [1.13, 2.95] | 1.82 [1.11, 2.98] | 1.05 [0.87, 1.28] | 1.05 [0.86, 1.29] |
| Quite frequently/Nearly always<sup>N</sup> | 2.00 [1.03, 3.91] | 2.21 [1.13, 4.36] | 1.07 [0.78, 1.47] | 1.08 [0.86, 1.29] |
| ≥ 20 weekly hours of care (binary) | 1.15 [0.80, 1.64] | 0.43 [0.15, 1.20] | 1.05 [0.75, 1.46] | 0.97 [0.68, 1.40] |
| Length of care (years) | 1.50 [0.97, 2.32] | 1.33 [0.82, 2.15] | 0.98 [0.81, 1.18] | 0.96 [0.78, 1.19] |
| Self-reported main caregiver | 1.11 [0.92, 1.35] | 1.24 [0.80, 1.93] | 1.04 [0.88, 1.24] | 1.06 [0.88, 1.27] |
| **PANEL 3: AGE 40 AND ABOVE** |  |  |  |  |
| CR expects you to care as the only dependable person (vs Never) |  |  |  |  |
| Rarely | 1.30 [1.02, 1.68] | 1.17 [0.89, 1.53] | 0.98 [0.61, 1.54] | 1.03 [0.63, 1.68] |
| Sometimes | 1.51 [1.16, 1.95] | 1.57 [1.20, 2.04] | 1.56 [1.92, 1.54] | 1.47 [0.87, 2.49] |
| Quite frequently | 1.11 [0.76, 1.60] | 1.11 [0.75, 1.63] | 1.23 [0.63, 2.42] | 1.06 [0.52, 2.14] |
| Nearly always | 1.08 [0.59, 1.96] | 1.10 [0.60, 2.07] | 0.81 [0.24, 2.65] | 0.58 [0.173, 1.95] |
| ≥ 20 weekly hours of care (binary) | 1.10 [0.74, 1.61] | 1.15 [0.76, 1.75] | 1.28 [0.61, 2.68] | 1.02 [0.47, 2.24] |
| Length of care (years) | 0.97 [0.84, 1.11] | 1.00 [0.87, 1.15] | 1.17 [0.92, 1.48] | 1.08 [0.84, 1.40] |
| Self-reported main caregiver | 0.93 [0.75, 1.16] | 0.92 [0.73, 1.18] | 1.56 [1.06, 2.29] | 1.52 [0.99, 2.35] |

Columns 1 and 3 contain results from four separate models per panel – each with one care variable. Columns 2 and 4 contain results from one model per panel containing all four care variables. All models additionally included as covariates: Age, sex, marital status and education. Full results from all models are provided as Supplementary Tables 1, 2 and 3.<sup>N</sup> Due to very few under 40 year olds reporting 'nearly always' (n=4) we merged these individuals into the 'quite frequently' group for these models. CR: care recipient.

The adjusted association between self-reported health and expectations of care was also inverse U-shaped, with the greatest health burden amongst those ‘rarely’ (odds ratio [OR] 1.36, 95%CI: 0.99-1.88) or ‘sometimes’ (OR: 1.40, 95%CI 0.97-2.01) expected to care compared to those never expected to care (Table 2). Self-reported main caregivers were more likely to self-report worse health, although this was also only seen amongst those aged ≥40 (OR: 1.52, 95%CI: 0.99-2.35). There was little association between providing 20 or more hours of care, or longer length of caregiving, and self-reported health. In multivariable models, poorer self-reported health and more chronic conditions were positively associated with age, lower educational attainment and lower workforce participation (Supplementary Table 1). Women had slightly higher counts of chronic conditions in these models, especially under age 40, and being previously but not currently married (partnered) was also associated with poorer health outcomes.

## DISCUSSION

Chronic conditions, often multiple morbidity, is common in South Africa and is concentrated amongst those who are socioeconomically disadvantaged, including those with physically and emotionally demanding roles. In this cross-sectional study of over 1000 people who cared for older adults living with or at risk of cognitive impairment in rural South Africa, we found a high prevalence of ill-health amongst caregivers, with almost half having one or more health conditions – despite having a median age of under 40. However, despite this high prevalence, the intensity of caregiving roles was not clearly or consistently associated with either numbers of reported chronic conditions or self-reported health. The notable exceptions were age-specific: older caregivers had an inverse U-shaped relationship such that the most caregiving was provided by those with intermediate levels of ill-health, and worse self-reported health amongst those who self-reported being main caregivers. Among those aged under 40, those with more chronic conditions were expected to act as sole caregivers more often.

Our headline findings, showing few significant associations between caregiving and caregiver health, and notably finding no significant associations for weekly hours of care, are contrary to studies in HICs (25). While it is hard to unpack the bi-directional association between ill-health and caregiving, especially in a cross-sectional study such as ours, this discrepancy may reflect important aspects of our rural LMIC context. Centrally, this rural setting is typified by intergenerational households, extended kinship connections and high levels of community connection. As we have previously highlighted (39), caregiving responsibilities in these households are widely shared among family members and community networks, as is common in LMIC settings (20, 47–49). This collective approach to caregiving contrasts with the often isolated caregiving experiences reported in HICs, where nuclear families may not have the same level of communal support (18, 25), and may explain our findings in three ways.

First, the potential of multiple caregivers may alleviate the stress and physical demands on any one single caregiver. This stress-reduction may be bolstered by societal expectations that caregiving is a shared social responsibility and a natural part of familial duties, which could mitigate the stress associated with caregiving (47, 49). Second, potential caregivers who are already unwell may be able to reduce their support as others are able to cover for them. Third, we have previously noted that care provision in this setting appears to be largely one of convenience, i.e., those family and friends who are locally resident and not otherwise occupied are called upon to care (39). As a result, the health of those called on to provide care may, in fact, be worse than non-carers if their availability is a function of ill-health. Unpicking which of these processes is occurring – and whether some combination of them might explain the U-shaped association seen in our data – is beyond this analysis and would require an in-depth examination of longitudinal qualitative data. But these contextual differences highlight the importance of considering cultural and social factors when examining the health impacts of caregiving across different settings.

Our observation that the greatest health burden was among those who were ‘sometimes’ expected to be the only dependable caregiver may reflect heightened stress due to intermittent and unpredictable caregiving demands, which can exacerbate existing health conditions (50, 51). Those in intermittent caring roles do not know when or how intensely they will be called upon to help, which disrupts routines and increases anxiety (22). This constant state of alert may cause emotional exhaustion and exacerbate both mental and physical health conditions (52). These caregivers experience a ‘dual burden’, balancing other life responsibilities while facing sudden caregiving demands, leading to increased stress and guilt (53). This precarious balance, combined with unpredictability, may trigger a stress response that weakens immune function and heightens the risk of stress-related illnesses such as cardiovascular disease and depression (51). Additionally, these caregivers may not receive the same recognition or resources as full-time caregivers, increasing their emotional and physical burden (54).

One important pattern seen in our study is the strong negative association between hours of care provided and self-reported health, but almost no association between care hours and the number of chronic conditions reported. This potentially highlights caregiving’s emotional and physical toll at a sub-clinical level (55), in line with other studies showing that long extended caregiving leads to worse self-perceived health due to emotional, social, and physical strain, even in the absence of additional chronic conditions (22, 50). This caregiving toll often extends to emotional wellbeing with many caregivers experiencing ‘anticipatory grief’, including the loss both of their loved one and their own identity, time, and emotional stability (54). Insofar as caregivers are unable to step away from caregiving despite worsening health they may feel trapped, reinforcing a vicious cycle where their health declines further (52).

As noted above, associations differed by age of caregiver. The strong association of greater caregiver expectations and more chronic conditions among younger caregivers may reflect limited experience and resources to balance elder caregiving and other life responsibilities, which can intensify stress and health deterioration (28, 56). This dual role arises frequently for grandchildren in particular in Africa, due to the migration of parents for work or to the death of parent(s) (55, 56). The causal pathway may also run the other way: younger people with chronic health problems were less likely to be in the workforce, and so were more often available to provide care. Again, longitudinal work is vital to determine which processes drive the association.

Among older adults, the association between being a main caregiver and increased likelihood of self-reported adverse health may reflect the cumulative physical and psychological burdens of caregiving which older adults are less able to mitigate than younger peers (22). This may relate to increasing caregiving intensity, including physical stressors on caregivers, the need to juggle their own health and the requirements of caring for others, or to the demands of care impacting their health more than for younger individuals. There is also the potential for older main caregivers ≥40 to have multiple care roles, e.g., as primary family provider, giving grandchild care, and caring for other elders. These multiple roles are often placed on older adults in this setting due to their eligibility for old age grants which are a substantial proportion of the rural South African economy (57, 58). In contrast, younger caregivers may find caregiving more manageable, with resilience arising from better physical health, greater stamina, and more effective coping strategies, including a sense of purpose from active social lives that buffers against caregiving stress (50, 59).

Our findings showed that caregivers were more likely to report adverse self-reported health, implying the need for a comprehensive consideration of caregiver health, delivering support that addresses physical health, emotional grief, and the subjective experiences of caregivers; this is likely to include mental health services, grief counselling, and respite care. High-income country (HIC) studies have shown that interventions can improve caregivers’ health (9, 60, 61). However, further research on rural African caregivers is needed to determine how best to support their health as they support others. Given that much caregiving in these settings will remain informal for the foreseeable future, providing respite care and educational programs is critical. Respite care in this context could offer temporary relief through in-home services, community-based day centres, or short-term residential facilities, giving caregivers time to rest or manage personal responsibilities (62). Additionally, educational programs should enhance caregivers’ skills in managing health conditions, such as training in medication administration and chronic disease management (14). These programs should also focus on mental health support and strategies to manage burnout, particularly among caregivers who themselves may suffer from chronic health issues (63, 64). Financial support, such as stipends or subsidies, could alleviate the economic burden, further enabling caregivers to continue their essential roles without undue stress (65). Overall, these interventions are crucial to enhancing caregiver well-being while improving the quality of care provided in informal settings.

### Strengths and limitations

While this study provides a comprehensive evaluation of the health of caregivers for over 100 individuals at high risk of cognitive impairment in rural South Africa, it has some limitations. The cross-sectional design limited our ability to determine the direction of causality in several areas; longitudinal studies are urgently needed to establish temporal relationships between caregiving and health outcomes in this setting. The self-reported nature of the data may have introduced response bias and social desirability effects, such as that seen in the disparity between high levels of both good self-reported health and multiple chronic health conditions. This pattern has been widely documented as the ‘wellbeing paradox’ (66, 67), highlighting the complexity of objective and subjective health measures. We are further investigating discrepancies between quantitative survey data and qualitative observations in other work on this project, but future work to link these data to objective health measures would be valuable.

## Conclusion

As poorer rural populations age, informal home-based caregiving will grow, generating increased need to find ways to support caregivers, mitigate burden, and enhance resilience. In the context of the provision of care to older people assessed as at risk of dementia in a resource-limited setting, this study provided valuable insights into the health of caregivers in rural South Africa. We identified a high prevalence of multiple chronic conditions across ages and noted that these compounded over time. At the same time, it appeared that intensive caregiving was not always associated straightforwardly with worse health, in a setting where caregiving is typically shared widely, and poor health is common. Nevertheless, there is a need for future research to advance our understanding of caregiving dynamics and inform targeted interventions in poor rural areas to optimise caregiver support and foster resilient caregiving communities.

## Acknowledgements

The research on which this study is based is nested within the MRC/Wits Rural Public Health and Health Transitions Research Unit from the School of Public Health at the University of the Witwatersrand and is responsible for the Agincourt Health and Socio-Demographic Surveillance System. The Unit is a node of the South African Population Research Infrastructure Network (SAPRIN) and receives support from the Department of Science and Innovation, the University of the Witwatersrand, the Medical Research Council, South Africa, and previously The Wellcome Trust (058893/Z/99/A; 069683/Z/02/Z; 085477/Z/08/Z; 085477/B/08/Z). This research was funded in whole, or in part, by the Wellcome Trust [Grant numbers 210479/Z/18/Z]. For the purpose of open access, the author has applied a CC BY public copyright licence to any Author Accepted Manuscript version arising from this submission.

## Disclaimer

The funders had no role in study design, data collection and analysis, decision to publish, or preparation of the manuscript

## SUPPLEMENTARY MATERIAL

**Supplementary Table 1.** Multivariable regression showing predictors for health outcome.

|  | Number of chronic conditions |  |  | Self-reported quality of health |  |  |
| --- | --- | --- | --- | --- | --- | --- |
|  | Model 1:<br>ALL | Model 2:<br>Below 40 | Model 3:<br>40 and above | Model 1:<br>ALL | Model 2:<br>Below 40 | Model 3:<br>40 and above |
| CR expects you to care as the only dependable person (vs Never) |  |  |  |  |  |  |
| Rarely | 1.33[ 1.05, 1.68] | 1.81 [1.15, 2.84] | 1.17[0.89,1.53] | 1.36 [0.99, 1.88] | 1.08[0.90, 1.27] | 1.03[0.63, 1.68] |
| Sometimes | 1.50 [ 1.18, 1.92] | 1.82 [ 1.11, 2.98] | 1.57[1.20 ,2.04] | 1.40 [0.97, 2.01] | 1.05[0.86, 1.29] | 1.47[0.87, 2.49] |
| Quite frequently | 1.34 [ 0.96, 1.89] | 2.21 [1.13,4.36] | 1.11 [0.75, 1.63] | 1.10 [0.64, 1.91] | 1.08[0.86, 1.29] | 1.06[0.52, 2.14] |
| Nearly always | 1.19 [ 0.64, 2.25] | - | 1.10 [0.60, 2.07] | 0.79 [0.28, 2.20] | - | 0.58[0.173, 1.95] |
| Weekly hours of Care | 1.02[ 0.69, 1.49] | 0.43 [ 0.15,1.20] | 1.18 [0.56, 0.98] | 1.11 [0.62, 1.97] | 0.97[0.68, 1.40] | 1.02[0.47, 2.24] |
| Length of care (years) | 1.04 [ 0.90, 1.20] | 1.33 [0.82, 2.15] | 1.15[0.76, 1.75] | 1.08 [0.87, 1.35] | 0.96[0.78, 1.19] | 1.08 [ 0.84, 1.40] |
| Self-reported main caregiver | 1.04 [ 0.84,1.29] | 1.24 [0.80, 1.93] | 1.00[0.87, 1.15] | 1.44[1.06, 1.94] | 1.06[0.88,1.27] | 1.52[0.99, 2.35] |
| Caregiver age (vs19-39) |  |  |  |  |  |  |
| < 18 | 0.36[0.22, 0.59] | 0.35[0.19, 0.62] | - | 0.22[0.14, 0.37] | 0.71[0.57, 0.87] | - |
| 40-59 | 1.67[1.26, 2.21] | - | 0.74[0.56, 0.98] | 1.83[1.26, 2.69] | - | 0.39 [0.22, 0.66] |
| ≥ 60 | 2.02 [1.40, 2.92] | - | - | 3.92[2.20, 6.97] | - | - |
| Sex (vs Male) |  |  |  |  |  |  |
| Female | 1.41 [1.15, 1.72] | 1.91[1.28, 2.82] | 1.23 [0.97, 1.54] | 1.14 [ 0.87, 1.49] | 1.09[0.93, 1.26] | 0.79 [0.53, 1.17] |
| Marital status (vs Married) |  |  |  |  |  |  |
| Never married | 0.83 [ 0.63, 1.10] | 0.92 [0.61, 1.38] | 1.01[0.62, 1.52] | 1.12 [0.77, 1.61] | 1.01[0.89, 1.20] | 1.68 [0.82, 3.43] |
| Previously married | 1.05 [ 0.85, 1.30] | 2.05[1.01, 4.17] | 1.00 [0.80, 1.26] | 1.28 [0.90, 1.81] | 1.22 [ 0.79, 1.23] | 1.24 [0.84, 1.84] |
| Work status (vs full-time work) |  |  |  |  |  |  |
| Part-time work | 1.24[ 0.86, 1.78] | 1.67[0.79, 3.59] | 1.06[0.69, 1.63] | 0.68 [0.42, 1.12] | 1.01[ 0.74, 1.35] | 0.54 [0.27, 1.08] |
| Seeking work | 1.1 8[0.86, 1.61] | 1.43[0.78, 2.62] | 1.08[0.74, 1.58] | 0.79 [0.53, 1.17] | 0.98[0.86, 1.61] | 0.94 [0.51, 1.69] |
| Out of workforce | 1.61 [1.21, 2.14] | 2.13[1.19, 3.80] | 1.24[0.87, 1.74] | 1.94 [1.30, 2.91] | 1.26 [0.98, 1.55] | 1.50 [0.84,2.67] |
| Education (vs Primary) |  |  |  |  |  |  |
| No Education | 0.89 [0.68, 1.16] | 0.87[0.09, 8.38] | 0.80 [0.63, 1.04] | 1.09 [0.68, 1.74] | 0.51 [0.12, 2.09] | 1.26 [0.76, 2.08] |
| Secondary Education | 0.91 [0.70, 1.17] | 1.02[0.57, 1.80] | 0.77 [ 0.57, 1.02] | 0.91 [0.63, 1.32] | 0.93 [0.74, 1.17] | 0.90[0.55, 1.47] |
| Tertiary Education | 0.60 [ 0.39, 0.93] | 0.49[0.19, 1.23] | 0.68 [0.42, 1.11] | 0.37 [0.22, 0.63] | 0.80 [ 0.58, 1.10] | 0.30[0.14, 0.62] |

**Supplementary Table 2.** Chronic conditions and adjusting for sociodemographic factors.

|  | Number of chronic conditions |  |  |  |  |
| --- | --- | --- | --- | --- | --- |
|  | Model 1 | Model 2 | Model 3 | Model 4 | Model 5 |
| CR expects you to care as the only dependable person (vs Never) |  |  |  |  |  |
| Rarely | - | - | - | 1.34 [1.07, 1.68] | 1.33[ 1.05, 1.68] |
| Sometimes | - | - | - | 1.49 [1.17, 1.88] | 1.50 [ 1.18, 1.92] |
| Quite frequently | - | - | - | 1.33[0.96, 1.84] | 1.34 [ 0.96, 1.89] |
| Nearly always | - | - | - | 1.20 [0.65, 2.20] | 1.19 [ 0.64, 2.25] |
| Weekly hours of Care | - | - | 1.18 [0.83, 1.67] | - | 1.02[ 0.69, 1.49] |
| Length of care (years) | - | 1.07 [0.93, 1.22] | - | - | 1.04 [ 0.90, 1.20] |
| Self-reported main caregiver | 1.11 [0.91, 1.36] | - | - | - | 1.04 [ 0.84, 1.29] |
| Caregiver age (vs19-39) |  |  |  |  |  |
| < 18 | 0.36[0.22, 0.59] | 0.37[0.23, 0.59] | 0.36[ 0.23, 0.58] | 0.36[0.23, 0.58] | 0.36[0.22, 0.59] |
| 40-59 | 1.67[1.28, 2.21] | 1.69[1.29, 2.22] | 1.71[1.31, 2.24] | 1.72[ 1.32, 2.25] | 1.67[1.26, 2.21] |
| ≥ 60 | 1.97[1.37, 2.84] | 1.96 [1.38, 2.81] | 2.00 [1.40, 2.85] | 2.08[ 1.46, 2.97] | 2.02 [1.40, 2.92] |
| Sex (vs Male) |  |  |  |  |  |
| Female | 1.39 [1.142, 1.70] | 1.42 [1.17, 1.72] | 1.42 [1.17, 1.72] | 1.45 [1.19, 1.75] | 1.41 [1.15, 1.72] |
| Marital status (vs Married) |  |  |  |  |  |
| Never married | 0.84 [0.63, 1.10] | 0.84 [0.64, 1.11] | 0.83 [0.63, 1.09] | 0.85 [0.64, 1.11] | 0.83 [ 0.63, 1.10] |
| Previously married | 1.02 [0.83, 1.26] | 1.02 [0.83, 1.25] | 1.03 [0.84, 1.26] | 1.02 [0.83, 1.25] | 1.05 [ 0.85, 1.30] |
| Work status (vs full-time work) |  |  |  |  |  |
| Part-time work | 1.16 [ 0.81, 1.69] | 1.12 [0.79, 1.60] | 1. 20[0.84, 1.71] | 1.12 [0.78, 1.59] | 1.24[ 0.86, 1.78] |
| Seeking work | 1.07 [0.78, 1.47] | 1.04 [0.77, 1.42] | 1.11 [0.82, 1.50] | 1.06 [0.78, 1.44] | 1.1 8[0.86, 1.61] |
| Out of workforce | 1.51 [1.13, 2.02] | 1.48 [1.12, 1.96] | 1.60 [ 1.22, 2.11] | 1.47 [1.11, 1.94] | 1.61 [1.21, 2.14] |
| Education (vs Primary) |  |  |  |  |  |
| No Education | 0.89 [0.68, 1.16] | 0.89[0.69, 1.14] | 0.89 [0.69, 1.15] | 0.91 [0.70, 1.17] | 0.89 [0.68, 1.16] |
| Secondary Education | 0.90 [0.70, 1.16] | 0.90 [0.71, 1.15] | 0.90 [ 0.71, 1.15] | 0.91[0.71, 1.16] | 0.91 [0.70, 1.17] |
| Tertiary Education | 0.59 [0.39, 0.91] | 0.61 [0.40, 0.92] | 0.62 [0.41, 0.94] | 0.63 [0.42, 0.95] | 0.60 [ 0.39, 0.93] |

**Supplementary Table 3.** Self-reported health and adjusting for sociodemographic factors.

|  | Self-reported quality of health |  |  |  |  |
| --- | --- | --- | --- | --- | --- |
|  | Model 1 | Model 2 | Model 3 | Model 4 | Model 5 |
| CR expects you to care as the only dependable person (vs Never) |  |  |  |  |  |
| Rarely | - | - | - | 1.28 [0.94, 1.75] | 1.36 [0.99, 1.88] |
| Sometimes | - | - | - | 1.47 [1.04, 2.10] | 1.40 [0.97, 2.01] |
| Quite frequently | - | - | - | 1.21 [0.71, 2.06] | 1.10 [0.64, 1.91] |
| Nearly always | - | - | - | 1.03 [0.38, 2.83] | 0.79 [0.28, 2.20] |
| Weekly hours of Care | - | - | 1.41 [0.82, 2.42] | - | 1.11 [0.62, 1.97] |
| Length of care (years) | - | 1.16 [0.95, 1.42] | - | - | 1.08 [0.87, 1.35] |
| Self-reported main caregiver | 1.46 [1.11, 1.92] | - | - | - | 1.44[1.06, 1.94] |
| Caregiver age (vs < 19-39) |  |  |  |  |  |
| Under 18 | 0.21[0.13, 0.36] | 0.21[0.13, 0.34] | 0.20 [0.13, 0.34] | 0.21 [0.13, 0.34] | 0.22[0.14, 0.37] |
| 40-59 | 1.81[1.25, 2.64] | 1.78[1.23, 2.58] | 1.85[ 1.28, 2.67] | 1.87 [1.29, 2.71] | 1.83[1.26, 2.69] |
| ≥ 60 | 3.76[2.13, 6.62] | 3.43[1.96, 6.02] | 3.62[2.08, 6.31] | 3.88 [2.21, 6.78] | 3.92[2.20, 6.97] |
| Sex (vs Male) |  |  |  |  |  |
| Female | 1.14[0.86, 1.48] | 1.19 [0.92,1.53] | 1.18 [0.91, 1.53] | 1.21 [0.94, 1.57] | 1.14 [ 0.87, 1.49] |
| Marital status (vs Married) |  |  |  |  |  |
| Never married | 1.09 [0.75, 1.58] | 1.08 [0.76, 1.56] | 1.07 [0.75, 1.54] | 1.10 [0.77, 1.58] | 1.12 [0.77, 1.61] |
| Previously married | 1.25[0.88, 1.77] | 1.22[0.87, 1.71] | 1.19 [0.85, 1.68] | 1.20 [0.86,1.69] | 1.28 [0.90, 1.81] |
| Work status (vs full-time work) |  |  |  |  |  |
| Part-time work | 0.67 [0.41, 1.11] | 0.67 [0.42, 1.09] | 0.68 [0.42, 1.10] | 0.67 [0.41, 1.10] | 0.68 [0.42, 1.12] |
| Seeking work | 0.76 [0.52, 1.12] | 0.74 [0.51, 1.09] | 0.72 [0.49, 1.07] | 0.77 [0.52, 1.13] | 0.79 [0.53, 1.17] |
| Out of workforce | 2.01 [1.34, 3.00] | 2.00 [1.35, 2.95] | 2.00[1.35, 2.95] | 1.88 [1.28, 2.79] | 1.94 [1.30, 2.91] |
| Education (vs Primary) |  |  |  |  |  |
| No Education | 1.05 [0.66, 1.67] | 1.10 [0.69, 1.74] | 1.10 [0.69, 1.74] | 1.19[0.75, 1.89] | 1.09 [0.68, 1.74] |
| Secondary Education | 0.91[0.63,1.30] | 0.92 [0.64, 1.31] | 0.92 [0.64, 1.32] | 0.93 [0.65, 1.33] | 0.91 [0.63, 1.32] |
| Tertiary Education | 0.36 [0.21, 0.61] | 0.38 [0.23, 0.65] | 0.40 [0.24, 0.68] | 0.41 [0.24, 0.70] | 0.37 [0.22, 0.63] |

